# Determinants of Qigong, Tai Chi, and Yoga Use for Health Conditions: A Systematic Review Protocol

**DOI:** 10.1101/2025.03.18.25324204

**Authors:** Ryan S. Wexler, Christopher Joyce, Rocky Reichman, Cora Pereira, Emma Fanuele, Emily Hurstak, Lance Laird, Helen Lavretsky, Chenchen Wang, Rob Saper, Karen S. Alcorn, Brian S. Mittman, Eric J. Roseen

**Author notes:** Corresponding Author: Ryan S. Wexler; Helfgott Research Institute, 2220 SW 1^st^ Ave, Portland, OR 97201.

## Abstract

**INTRODUCTION:** Mind-body movement interventions such as qi gong, tai chi, and yoga are recommended in clinical practice guidelines to improve outcomes for several health conditions. However, use of these interventions for health conditions, or the integration of these interventions within healthcare settings, is low. A systematic synthesis of implementation determinants (i.e., barriers and facilitators) is needed to increase adoption. Similarly, determinants may influence other implementation outcomes, such as scalability or sustainability of these interventions in a healthcare system or community organization. Thus, in conducting this review we aim to identify determinants of qi gong, tai chi, and yoga use for health conditions. The secondary aim is to evaluate whether barriers and facilitators differ by intervention type, health condition, implementation setting, or implementation outcome.

**METHODS AND ANALYSIS:** We conducted a comprehensive search of electronic databases (MEDLINE, EMBASE, Web of Science, CINAHL, PsycInfo) through May 2024 and a grey literature search (Google Scholar, Cochrane Central Register of Controlled Trials, ClinicalTrials.gov, the WHO Clinical Trials database) through March 2025. We will include original research articles in English that identify barriers and facilitators to adoption of qi gong, tai chi, and yoga by adults with health conditions. Study quality will be assessed using the Mixed Methods Appraisal Tool. We will code each article using a codebook informed by the Consolidated Framework for Implementation Research (CFIR), a comprehensive taxonomy of implementation determinants. Findings will be presented as a narrative synthesis. We will report on how barriers and facilitators may relate to intervention type (qi gong, tai chi, yoga), health condition (e.g., low back pain, fall prevention), implementation settings (e.g., primary care clinic, community organization) or implementation outcome (e.g., adoption, sustainability).

**ETHICS AND DISSEMINATION:** Ethics approval will not be obtained for this review of published, publicly accessible data. The results from this systematic review will be disseminated through conference presentations and journal publications.

**STRENGTHS AND LIMITATIONS OF THIS STUDY:**

- There are no prior comprehensive reviews of the determinants (i.e., barriers and facilitators) of qi gong, tai chi and yoga use. This review will examine whether barriers and/or facilitators vary by type of mind-body movement intervention, health condition, implementation setting, or implementation outcome.
- Our search has been guided by librarians with expertise in systematic review methodology and informed by the PEER Review of Electronic Search Strategies (PRESS) guidelines. Reporting of results will be informed by the PRISMA-P checklist for systematic reviews.
- We will use the updated version of the Consolidated Framework for Implementation Research (CFIR, version 2.0) to report implementation factors using consistent concepts to facilitate use of the review by researchers, healthcare systems, and community organizations. Our findings will support the development of implementation strategies to increase the adoption of qi gong, tai chi, and yoga for health conditions.
- Due to the complex nature of describing implementation determinants and characterizing them within the Consolidated Framework for Implementation Research, our review is limited to papers written in English which may restrict the generalizability of our implementation findings to predominantly English-speaking healthcare settings.

## INTRODUCTION

Qi gong, tai chi, and yoga are mind-body movement interventions that combine body positions or postures with mindful movement, controlled breathing, and meditation.^1–5^ The practice of these mind-body movement interventions can lead to improved physical function, general health and better disease-specific outcomes.^6–13^ For example, tai chi has been shown to improve balance and prevent falls, increase function in older adults with knee and hip osteoarthritis, and improve several patient reported outcomes among adults with neuropsychiatric and cognitive disorders.^6–8,14–17^ Similarly, yoga has been shown to improve outcomes for adults with low back pain, diabetes and cancer-related symptoms.^18–22^ Accordingly, qi gong, tai chi, and yoga are recommended in major clinical practice guidelines for several common adult health conditions as shown in **Table 1**.^23–30^ While it is estimated that 20% of adults in the United States practice these mind-body movement interventions,^31^ and use of yoga has exponentially increased over time,^32,33^ use is predominantly focused on mind-body movement practice as a form of exercise to support general wellbeing.^34^ Thus, despite growing evidence that qi gong, tai chi, and yoga can improve outcomes and reduce morbidity in persons living with common health conditions,^9,23–30,35^ utilization of these interventions for specific health conditions remains low.^34^

**Table 1.** Select clinical practice guidelines that recommend yoga, tai chi, and qi gong for specific health conditions.

| Organization | Recommendation |
| --- | --- |
| American College of Physicians | **Yoga/Tai Chi** recommended for chronic low back pain in 2017 guideline<sup>26</sup> |
| American College of Rheumatology | **Tai Chi** recommended for hip and knee osteoarthritis in 2019 guideline<sup>25,27</sup><br>**Qi Gong** recommended for rheumatoid arthritis<sup>30</sup> |
| American Society for Clinical Oncology/ Society of Integrative Oncology | **Yoga/Tai Chi/Qi Gong** recommended during cancer care for symptom management in 2022 guideline<sup>28</sup> |
| International Society on Hypertension | **Yoga** recommended for hypertension in 2020 guideline<sup>23</sup> |
| American Diabetes Association | **Yoga/Tai Chi** recommended for diabetes management in 2024 guideline<sup>29</sup> |
| US Preventative Services Task Force | **Tai Chi** recommended for falls prevention in 2024 guideline<sup>24</sup> |

In the healthcare context, implementation science involves the study of methods to promote the systematic uptake of evidence-based practices into routine clinical care.^36^ Understanding the determinants (i.e., barriers and facilitators) of evidence-based clinical interventions is an important first step in developing effective implementation strategies that increase the use of these interventions. Furthermore, barriers to intervention use can occur at multiple socio-ecological levels.^37–42^ For example, barriers of mind-body movement interventions in healthcare settings can occur at the level of the patient (e.g., high out-of-pocket costs), referring provider (e.g., low self-efficacy to describe or refer to tai chi), health system technology (e.g., referral to yoga not possible in electronic health record), or policy-maker (e.g., payers not reimbursing mind-body movement interventions).^43–49^ Thus, identifying and prioritizing modifiable barriers is needed to design tailored implementation strategies that can increase the use of qi gong, tai chi and yoga for specific health conditions, as recommended by clinical practice guidelines.^50^

The Consolidated Framework for Implementation Research (CFIR) was developed to guide the comprehensive and systematic assessment of implementation determinants.^51^ CFIR includes 67 distinct constructs–providing a comprehensive taxonomy of implementation determinants–which are organized into five domains: Innovation (e.g., tai chi), Outer Setting (e.g., policy makers including insurers), Inner Setting (e.g., a health system or clinic), Individuals (e.g., clinicians that may refer to Qi Gong), and the Implementation Process.^51^ The systematic assessment and mapping of determinants to these specific constructs can be used to develop tailored strategies to increase the use of evidence-based practices.^52^ As clinical guidelines increasingly support the use of qi gong, tai chi, and yoga for specific health conditions, and healthcare systems or community organizations begin to implement these interventions,^53–56^ there is a growing need to understand implementation determinants to increase the effectiveness and efficiency of these implementation efforts.^47,57–59^ While the explicit use of implementation science theories or frameworks, like CFIR in implementation of mind-body movement interventions is just emerging,^47,60^ there are lessons to learn from prior literature. However, we are unaware of systematic reviews that summarize the existing knowledge around implementation determinants of mind-body movement interventions.

One reason for the absence of prior reviews may be the limited familiarity, awareness, or use of CFIR (or other evolving determinant frameworks^42,61^) within the clinical research communities studying mind-body movement interventions. Yet, applying the common language codified in CFIR to code the existing literature may help synthesize and better document prior work, thus elucidating known barriers and informing future implementation efforts. It may also be that heterogeneity exists in the factors that influence the implementation of mind-body movement interventions (e.g., if barriers to implementing tai chi for knee osteoarthritis are different than the barriers that impede implementation of yoga in cancer care). Understanding these differences will aid the development of implementation strategies that are appropriately tailored to the intervention, health condition, implementation setting, or the relevant implementation outcomes.

The purpose of this systematic review will be to: 1) apply CFIR constructs to identify and report barriers and facilitators to the use of qi gong, tai chi, and yoga in treating various health conditions, and 2) identify sources of heterogeneity in determinants of intervention use, including variation in barriers and facilitators by type of mind-body movement intervention, health condition, implementation setting, or implementation outcome.^62^

## METHODS

### Design

We will conduct a systematic review with a qualitative meta-analysis of original studies to synthesize evidence on implementation barriers and facilitators.^62^ To organize and present our findings in common language that subsequently informs the development of implementation strategies, we will use an *a priori* codebook, informed by CFIR 2.0, to code and organize data from the original studies.^51^ During data extraction, we will code data from the included primary studies in order to organize all information into the CFIR domains and constructs (see **Table 2**).^62^ We will also allow for emerging themes not captured by the CFIR constructs.

**Table 2.** Overview of Consolidated Framework for Implementation Research (CFIR) domains.

| Domain | Description |
| --- | --- |
| Innovation | The innovation is the “thing” being implement (i.e., qi gong, tai chi, or yoga for specific health conditions). Characteristics of the innovation (e.g., innovation cost, evidence base for a specific health condition, relative advantage versus other already available innovations) will be characterized as barriers or facilitators. |
| Individuals | The roles and characteristics of individuals. We anticipate that the main ‘individuals involved’ will be the innovation recipients (i.e., patients who receive mind-body movement interventions) and intervention deliverers (instructors, clinicians who refer) and health system leaders. We will categorize papers as reflecting patient perspectives or non-patient perspectives or both. For these groups we will identify barriers and facilitators align with the needs, capability, motivation, and opportunity of these various individuals to use or implement recommend mind-body movement interventions for health conditions. |
| Inner Setting | The setting in which the innovation is implemented, e.g., primary care clinic or community organization. There may be multiple Inner Settings and/or multiple levels within the Inner Setting, e.g., hospital, department, clinic. Characteristics of the inner setting (e.g., physical space to practice tai chi, information technology infrastructure to facilitate referrals to qi gong, communication channels between primary care providers and yoga instructors) will be characterized as barriers or facilitators. |
| Outer Setting | The setting in which the Inner Setting exists (e.g., surrounding communities) or other external pressures (e.g., state or federal policy, insurance). Characteristics of the outer setting (e.g., local attitudes toward mind-body movement interventions, local conditions available to support implementation, policies or laws, or external pressure to implement mind-body movement interventions) will be characterized as barriers or facilitators. |
| Process | The activities and strategies used to implement the innovation. This involves documentation of the implementation process and what activities or strategies were used to implement the innovation. For studies that have defined implementation efforts, characteristics of the implementation process (e.g., forming teams to implement mind-body movement interventions, compacity to assess needs of inner setting or to tailor implementation process) will be characterized as barriers or facilitators. |

The protocol for this systematic review was registered with the international prospective register of systematic reviews (PROSPERO #CRD42024569493). Reporting of the review findings will be informed by the PRISMA guidelines study protocols (PRISMA-P).^63^

### Eligibility Criteria

This systematic review will include original peer-reviewed empirical studies including cross-sectional and longitudinal observational studies, randomized and non-randomized clinical trials, and qualitative and mixed method evaluations of implementation or quality improvement programs. We will conduct reference list checking, as described by the Terminology, Application, and Reporting of Citation Search (TARCiS) statement,^64^ from conference proceedings, commentaries, editorials, dissertations, letters, books, book chapters, and reviews, but will exclude these types of articles. Relevant systematic reviews, commentaries, and literature reviews will also be searched for original research references, but excluded from full text review and extraction

#### Inclusion Criteria

We will include qualitative or quantitative studies that examine determinants (barriers and facilitators) of implementation outcomes for qi gong, tai chi, or yoga in adults with health conditions. We will include studies of adults with a specific health condition or those considered “at-risk” for a given health condition and described by the authors as such (e.g., older adults at risk for falls, cancer survivors). We will also include studies of other key informants (e.g., clinicians, health system or community leaders, policy makers). For example, we will include studies of clinicians if the mind-body movement intervention is relevant to a health condition they manage (e.g., geriatricians may recommend tai chi for knee osteoarthritis or fall prevention). We will also include multicomponent interventions (e.g., combined acupuncture-yoga intervention, mindfulness-based stress reduction) if we can identify facilitators or barriers specific to the qi gong, tai chi, or yoga component of the program.^60,65^ Clinical trials will be included if they provide data on barriers and facilitators to mind-body movement intervention use thought to be relevant outside the context of the clinical trial. For example, we would include a study of yoga which measures feasibility and gathers information about barriers and facilitators to practicing yoga via semi-structured interviews or questionnaires. By contrast, we will exclude clinical trials that only measure implementation outcomes (e.g., measuring feasibility without collecting information on barriers or facilitators) or studies identifying barriers or facilitators that may only be relevant to the conduct of clinical trials (e.g., barriers to recruitment of trial participants).

#### Exclusion Criteria

Healthy populations (i.e., those without a specified health condition or not designated as at-risk for a health condition); populations under the age of 18 years; conference proceedings, commentaries, editorials, dissertations, letters, books, book chapters, and reviews; and articles written in languages other than English will be excluded. We will exclude studies written in languages other than English because it may be difficult to accurately code translated text while maintaining a common language from the field of implementation science (i.e., CFIR constructs) where term homonymity, synonymy, and instability are major challenges.^66–68^

### Search Strategy

A comprehensive search of studies was conducted from database inception until May 2024 using the electronic databases: MEDLINE (OVID platform), Embase (OVID platform), CINAHL, PsycInfo, and Web of Science. In addition, a grey literature search was conducted using Google Scholar, the Cochrane Central Register of Controlled Trials, ClinicalTrials.gov, and the WHO International Clinical Trials database in March of 2025. Search terms were variations on two concepts: qi gong, tai chi, or yoga AND barrier, facilitators, or implementation. Search strings varied slightly depending on the MESH and subject heading terms within each database.

Truncation (*) was used to capture variations in terms. Results were limited to the English language. See **Appendix 1** for full search strategies for each database. Endnote was used to remove duplicates and double-checked by librarians with expertise in systematic review methods. Additional duplicates were identified and removed in the systematic review software, Covidence (Veritas Health Innovation, Melbourne, Australia).

### Data Extraction

Through team meetings and the use of exemplar articles, an extraction form was developed **(Appendix 2)** to capture information on each study. We will extract the following information on general study characteristics: author(s), year, country where the study was conducted, study design, sample size, and data collection method (e.g., interviews, focus groups, surveys). We will collect information on the sample including participant demographic characteristic such as age, sex, race/ethnicity, and other characteristics that define the population of interest (e.g., military veterans, healthcare workers). We will identify what stakeholder perspectives(s) are being captured (e.g. patients, instructors of mind-body movement interventions, primary care providers, medical specialists, medical educators, hospital leadership, community leaders, policy makers). We will extract information on strength of evidence of the intervention for the specific health condition as justified by the original investigator. The authors rationale for implementation efforts will be characterized as: 1) the mind-body movement intervention is recommended for the specific health condition (e.g., a guideline recommending yoga for low back pain^9^); 2) a component of the mind-body movement intervention is recommended for the specific health condition (e.g., a guideline recommending exercise or mindfulness for cardiovascular health^69^); or 3) implementation is occurring without a clinical practice guideline supporting either the intervention or its components for the specific health condition.

We will extract information on several potential sources of heterogeneity in our evaluation of implementation determinants for mind-body movement intervention use for health conditions. First, we will identify the intervention type (i.e., qi gong, tai chi, yoga) and intervention delivery method (e.g., in-person, virtual). Second, we will identify health conditions of interest (e.g., low back pain, knee osteoarthritis). Third, the implementation setting will be characterized by the location where implementation is occurring (e.g., primary care clinic, community organization). We will also document if practice of qi gong, yoga, or tai chi occurs elsewhere (e.g., if primary care providers refer to community-based interventions or when virtually delivered interventions are practiced at home). When possible, we will also identify the specific implementation outcome(s) of interest in each study, with implementation outcomes defined by Proctor *et al* shown in **Table 3**.^70^ This will allow us to evaluate whether determinants vary by implementation outcome (e.g., was the determinant a barrier to adoption and/or sustainability?). Acceptability, appropriateness, and feasibility are considered pre-implementation outcomes, i.e., outcomes that can precede implementation efforts that can predict other implementation outcomes such as adoption or sustainability.^71^

**Table 3.** Definitions of Implementation Outcomes. ^70^

| Implementation Outcome | Definition |
| --- | --- |
| Acceptability<sup>a</sup> | The perception among implementation stakeholders that a given treatment, service, practice, or innovation is agreeable, palatable, or satisfactory, e.g., do patients or providers think that mind-body movement interventions are attractive as a treatment option? |
| Adoption | The intention, initial decision, or action to try or employ an innovation or evidence-based practice. Adoption also may be referred to as “uptake.” For example, what proportion of patients with knee osteoarthritis are referred to tai chi within a primary care clinic. |
| Appropriateness<sup>a</sup> | The perceived fit, relevance, or compatibility of the innovation or evidence-based practice for a given practice setting, provider, or consumer; and/or perceived fit of the innovation to address a particular issue or problem, e.g., do patients, providers and health system leaders perceive tai chi as appropriate for preventing falls?. |
| Cost | The cost impact of an implementation effort. |
| Feasibility<sup>a</sup> | The extent to which a new treatment, or an innovation, can be successfully used or carried out within a given agency or setting (e.g., primary care clinic, community organization). |
| Fidelity | The degree to which an intervention or innovation was implemented as it was prescribed in the original protocol or as it was intended by the program developers, e.g., is tai chi delivered in real-world settings is similar to tai chi delivered in clinical trials that have informed clinical practice guidelines? |
| Penetration | The integration of a practice within a service setting and its subsystems, e.g., the number of clinics or community organization locations that offer yoga for low back pain. |
| Sustainability | The extent to which a newly implemented treatment is maintained or institutionalized within a service setting’s ongoing, stable operations. |
| <sup>a</sup>Acceptability, appropriateness, and feasibility are considered pre-implementation outcomes, i.e., outcomes that can precede implementation efforts that can predict other implementation outcomes such as adoption or sustainability.<sup>71</sup> |  |

Barriers and facilitators will be identified through coding of relevant quantitative data (e.g., surveys) and qualitative findings (e.g., themes) from the results section of each study using a codebook informed by CFIR version 2.0.^51^ We have piloted this codebook in several prior and ongoing studies designed to inform the development of implementation strategies.^47,50^ We will also allow for emerging determinants (i.e., those that are not captured by CFIR) to arise throughout the coding process. Two reviewers will independently extract data from each of the primary studies. Disagreements in data extraction will be resolved by senior reviewers and/or team consensus meetings.

### Data Synthesis and Qualitative Meta Analysis

We will describe the frequency of identified barriers and facilitators and use heatmaps to illustrate high-density areas of CFIR.^72^ We will explore whether barriers and facilitators differ based on intervention type (e.g., tai chi vs. yoga), health condition (e.g., back pain vs cancer symptom management), setting (e.g., primary care clinic vs. community organization), and implementation outcome (e.g., adoption vs sustainability). For health condition, we will explore whether barriers and facilitators differ when qi gong, tai chi, and yoga are being implemented for different clinical indications (e.g., fall prevention, osteoarthritis, or symptom management in cancer care). For setting, we will explore whether barriers/facilitators differ when qi gong, tai chi, and yoga are implemented in different contexts. Clinical settings may include primary care clinics, medical specialty clinics (e.g., pain medicine, rheumatology, orthopedics, physical medicine and rehabilitation, integrative medicine, sports medicine), community settings (e.g., nursing homes, community centers, parks, home-based programs), or other specific settings where qi gong, tai chi, or yoga may be delivered for health conditions. For implementation outcomes, we will explore whether barriers and facilitators differ when a specific implementation outcome is specified. We will characterize each study as having no specified implementation outcomes or one or more of the following implementation outcomes: acceptability, appropriateness, feasibility, adoption, fidelity, penetration, cost, and sustainability of intervention delivery.^70^

### Risk of Bias Assessment

Study quality will be assessed using the Mixed Methods Appraisal Tool (MMAT) Version 2018.^73^ The MMAT is a grading check list that can be used to assess the risk of bias in five types of study designs: 1) Qualitative, 2) Quantitative randomized controlled trials, 3) Quantitative non-randomized trials, 4) Quantitative descriptive, and 5) Mixed methods. Two reviewers will independently use the appropriate checklist to grade each included study. Discrepancies between the two reviewers will be discussed at team meetings and/or arbitrated by a third reviewer.

## DISCUSSION

The benefits of qi gong, tai chi, and yoga have been established for several health conditions. Thus, we anticipate our systematic review will identify studies on barriers and facilitators to mind-body movement intervention use for specific health conditions where there is consensus that these interventions are helpful. For example, guidelines endorse one or more of these mind-body movement interventions for low back pain (American College of Physicians^9^), osteoarthritis (American College of Rheumatology and the Osteoarthritis Research Society International guidelines^25^ and the American College of Rheumatology and Arthritis Foundation^74^), hypertension (International Society of Hypertension Global Hypertension^23^), and fall prevention (United States Preventive Services Task Force among exercise interventions^24^). While several studies have looked at barriers to implementation of a specific intervention or within a particular clinical context, we are unaware of any publications that have synthesized what is known about implementing qi gong, tai chi, or yoga across broad settings where people with health conditions receive services.

Identifying barriers is essential for developing effective implementation strategies that can support their adoption in routine care settings. Implementation strategies that target specific known barriers may be more effective than general strategies. While developing implementation strategies that have multiple components is also thought to be effective, it is important to tailor resources to those strategies most likely to be successful within a specific context.^75,76^ Thus, identifying a range of barriers and facilitators, and whether they are relevant to a range of contexts, is essential to inform future dissemination and implementation efforts in research and practice.^77,78^ Our qualitative meta-analysis will help us to organize and interpret information on implementation determinants using the common set of terminology defined in CFIR domains and constructs.

## Data Availability

All data produced in the present study are available upon reasonable request to the authors

## Patient and Public Involvement

Patients and the public will not be involved in the study design, data extraction, or interpretation of results.

## Ethics and Dissemination

Ethics approval will not be obtained for this review of published, publicly accessible data. The results from this systematic review will be disseminated through conference presentations and journal publications. Based on our preliminary search results, we anticipate a majority of the papers included in our review will reflect the patient perspective, i.e., patient-related barriers to use of mind-body interventions for health conditions. Due to this potential asymmetry in data on intervention barriers and facilitators among different stakeholder perspectives, we anticipate that findings from our systematic review and qualitative meta-analysis may be split into two reports, 1) patient perspectives and 2) provider, staff, and healthcare system leader perspectives. Additionally, this review will inform the implementation of tai chi in four large healthcare systems in the context of a multi-site embedded pragmatic trial funded by the National Center for Complementary and Integrative Health (NIH Grant #: UG3AT012413). We also anticipate our review will inform other implementation efforts of qi gong, tai chi and yoga for specific health conditions in health care systems or community organizations.

## Author Contributions

*Study concept and design:* RSW, CJ, RR, CP, and EJR

*Acquisition, analysis, or interpretation of data:* RSW, CJ, RR, CP, EM, EH HL, CW, RS, BM, KSA, JD, and EJR.

*Critical revision of the manuscript for important intellectual content:* RSW, CJ, RR, CP, EM, EH, HL, CW, RS, BM, KSA, JD, and EJR.

*Statistical analysis:* n/a

*Obtained funding:* n/a

*Administrative, technical, or material support: CJ, EJR*.

*Study supervision:* EJR

## Acknowledgements

We would like to acknowledge Joanne Doucette, an expert systematic review librarian who helped our team develop the search strategy for this review.

## Study Dates

Work on the protocol began in February 2024. Screening began after the protocol was registered on PROSPERO and as the protocol was submitted for peer review. It is expected the study will be completed by June 2025.

## Appendix 1. Detailed search strategy

**Database:** MEDLINE(R) ALL

**Host:** Ovid

**Data parameters:** 1946-05/18/2024

**Date of search:** 05/18/2024

**Results:** 4347

**Search Strategy:**

(Tai Ji/ OR Yoga/ OR Qigong/ OR (Tai Ji.tw. OR Tai-ji.tw. OR Tai Chi.tw. OR Taiji.tw. OR Taijiquan.tw. OR tai ji quan.tw. OR T’ai Chi.tw. OR tai chi chuan.tw. OR yoga.tw. OR yogic meditation.tw. OR qigong.tw. OR Qi Gong.tw. OR Ch’i kung.tw. OR Chi kung.tw.))

AND

(barrier*.tw. OR obstacle*.tw. OR challeng*.tw. OR difficult*.tw. OR issue*.tw. OR problem*.tw. OR imped*.tw. OR limit*.tw. OR struggle*.tw. OR complexit*.tw. OR disadvantage*.tw. OR restriction*.tw. OR facilitat*.tw. OR motivat*.tw. OR enabl*.tw. OR accessib*.tw. OR determin*.tw. OR implement*.tw.)

Limit to English

**Database:** Embase

**Host:** Ovid

**Data parameters:** 1974-05/19/2024

**Date of search:** 05/19/2024

**Results:** 5957

**Search Strategy:**

(Tai Chi/ OR Yoga/ OR Qigong/ OR (Tai Ji.tw. OR Tai-ji.tw. OR Tai Chi.tw. OR Taiji.tw. OR Taijiquan.tw. OR tai ji quan.tw. OR T’ai Chi.tw. OR tai chi chuan.tw. OR yoga.tw. OR yogic meditation.tw. OR qigong.tw. OR Qi Gong.tw. OR Ch’i kung.tw. OR Chi kung.tw.))

AND

(barrier*.tw. OR obstacle*.tw. OR challeng*.tw. OR difficult*.tw. OR issue*.tw. OR problem*.tw. OR imped*.tw. OR limit*.tw. OR struggle*.tw. OR complexit*.tw. OR disadvantage*.tw. OR restriction*.tw. OR facilitat*.tw. OR motivat*.tw. OR enabl*.tw. OR accessib*.tw. OR determin*.tw. OR implement*.tw.)

Limit to English

Omitted conference abstracts, books, and pre-prints

**Database:** CINAHL with Full Text

**Host:** EBSCOhost

**Data parameters:** 1981-05/18/2024

**Date of search:** 05/18/2024

**Results:** 2633

**Search Strategy:**

((MH “Tai Chi”) OR (MH “Yoga”) OR (MH “Qigong”) OR Tai Ji OR Tai-ji OR Tai Chi OR Taiji OR Taijiquan OR T’ai Chi OR yoga OR yogic meditation OR qigong OR Qi Gong OR Ch’i kung OR Chi kung))

AND

(barrier* OR obstacle* OR challeng* OR difficult* OR issue* OR problem* OR imped* OR limit* OR struggle* OR complexit* OR disadvantage* OR restriction* OR facilitat* OR motivat* OR enabl* OR accessib* OR determin* OR implement*)

Limit to English and Peer-review

**Database:** APA PsycInfo

**Host:** Ovid

**Data parameters:** 1806-05/18/2024

**Date of search:** 05/18/2024

**Results:** 1441

**Search Strategy:**

(Yoga/ OR (Tai Ji.tw. OR Tai-ji.tw. OR Tai Chi.tw. OR Taiji.tw. OR Taijiquan.tw. OR tai ji quan.tw. OR T’ai Chi.tw. OR tai chi chuan.tw. OR yoga.tw. OR yogic meditation.tw. OR qigong.tw. OR Qi Gong.tw. OR Ch’i kung.tw. OR Chi kung.tw.))

AND

(barrier*.tw. OR obstacle*.tw. OR challeng*.tw. OR difficult*.tw. OR issue*.tw. OR problem*.tw. OR imped*.tw. OR limit*.tw. OR struggle*.tw. OR complexit*.tw. OR disadvantage*.tw. OR restriction*.tw. OR facilitat*.tw. OR motivat*.tw. OR enabl*.tw. OR accessib*.tw. OR determin*.tw. OR implement*.tw.)

Limit to English, Peer-reviewed, no dissertations

**Database:** Web of Science

**Host:** Clarivate

**Data parameters:** Core Collection, 1900-05/18/2024

**Date of search:** 05/18/2024

**Results:** 2931

**Search Strategy:**

1. (TI=("Tai Ji" OR Tai-ji OR "Tai Chi" OR Taiji OR Taijiquan OR "T’ai Chi" OR yoga OR "yogic meditation" OR qigong OR "Qi Gong" OR "Ch’i kung" OR "Chi kung") OR AB=("Tai Ji" OR Tai-ji OR "Tai Chi" OR Taiji OR Taijiquan OR "T’ai Chi" OR yoga OR "yogic meditation" OR qigong OR "Qi Gong" OR "Ch’i kung" OR "Chi kung"))
2. (TI=(barrier* OR obstacle* OR challeng* OR difficult* OR issue* OR problem* OR imped* OR limit* OR struggle* OR complexit* OR disadvantage* OR restriction* OR facilitat* OR motivat* OR enabl* OR accessib* OR determin* OR implement*) OR AB=(barrier* OR obstacle* OR challeng* OR difficult* OR issue* OR problem* OR imped* OR limit* OR struggle* OR complexit* OR disadvantage* OR restriction* OR facilitat* OR motivat* OR enabl* OR accessib* OR determin* OR implement*))
3. #1 AND #2
4. #1 AND #2 and English (Languages)

Kept WOS records (omitted PDTA, Korean, SciELO, Medline)

**Database:** Google Scholar

**Host:** Google

**Data parameters:** N/A

**Date of search:** March 5, 2025

**Results:** The first 200 results were **used**

**Search Strategy:**

intitle:(tai chi OR qigong OR yoga) (barrier OR barriers OR facilitator OR facilitators)

**Database:** Cochrane Central Register of Controlled Trials

**Host:** Wiley

**Data parameters:** Issue 2 of 12, February 2025

**Date of search:** March 5, 2025

**Results:** 375

**Search Strategy:**

“tai chi” OR qigong OR yoga OR “Tai Ji” OR “Tai-ji” OR Taiji OR Taijiquan OR “T’ai Chi” OR “yogic meditation” OR “Qi Gong” OR “Ch’i kung” OR “Chi kung” in Title Abstract Keyword AND barrier OR facilitator in Title Abstract Keyword - (Word variations have been searched)

**Database:** ClinicalTrials.gov

**Host:** National Library of Medicine

**Data parameters:** 200-03/06/2025

**Date of search:** March 6, 2025

**Results:** 43

**Search Strategy:**

(tai chi OR qigong OR yoga OR Tai Ji OR Tai-ji OR Taiji OR Taijiquan OR T’ai Chi OR yogic meditation OR Qi Gong OR Ch’i kung OR Chi kung) AND (barrier OR barriers OR facilitator OR facilitators)

Limited to completed studies only

**Database:** WHO International Clinical Trials Database

**Host:** World Health Organization / International Clinical Trials Registry Platform (ICTRP)

**Data parameters:** 2006-03/06/2025

**Date of search:** March 6, 2025

**Results:** 20

**Search Strategy:**

(“tai chi” OR qigong OR yoga) AND (barrier* OR facilitator*)

## Appendix 2. Extraction forms

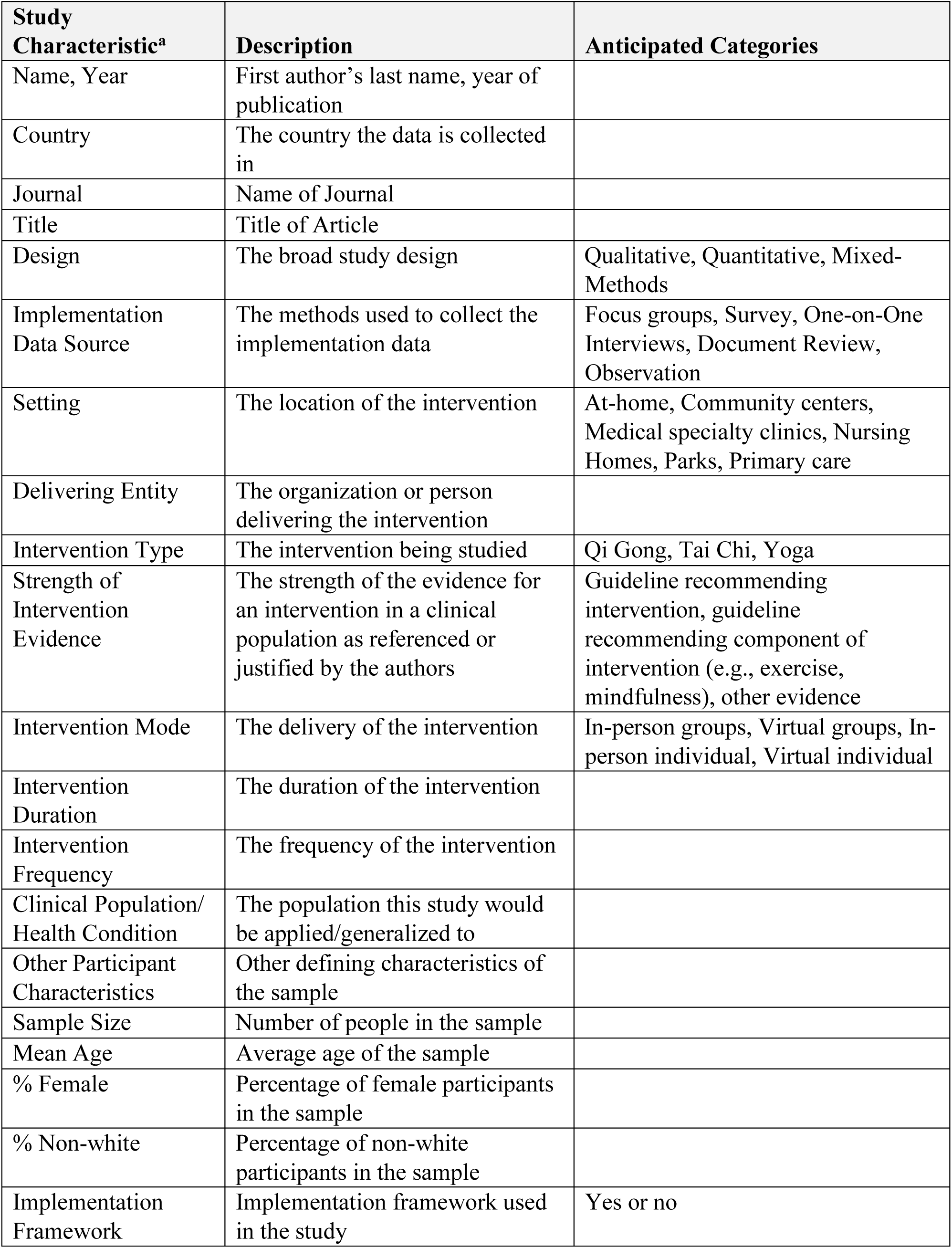
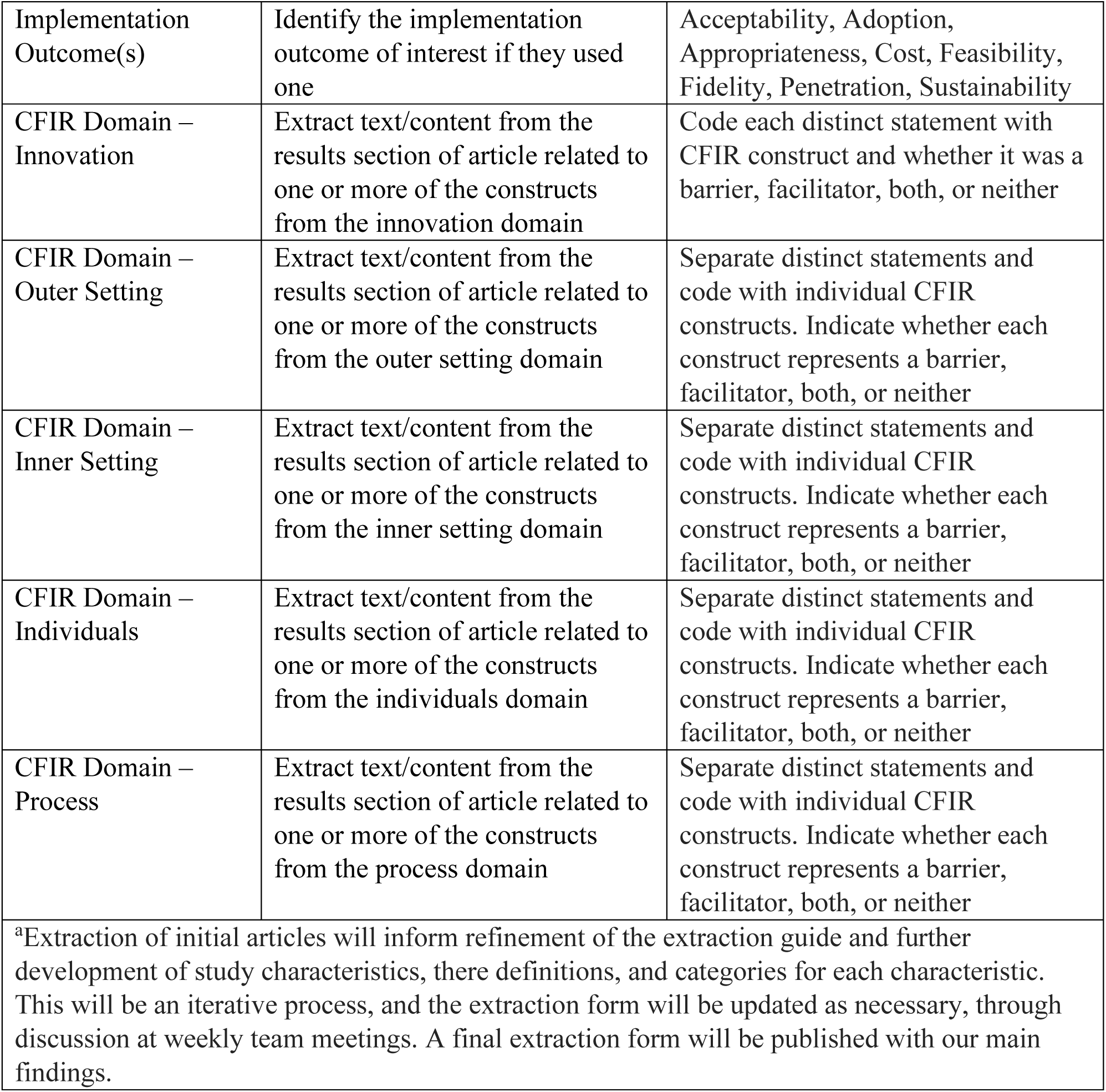

| **Study Characteristic<sup>a</sup>** | **Description** | **Anticipated Categories** |
| --- | --- | --- |
| Name, Year | First author's last name, year of publication |  |
| Country | The country the data is collected in |  |
| Journal | Name of Journal |  |
| Title | Title of Article |  |
| Design | The broad study design | Qualitative, Quantitative, Mixed-Methods |
| Implementation Data Source | The methods used to collect the implementation data | Focus groups, Survey, One-on-One Interviews, Document Review, Observation |
| Setting | The location of the intervention | At-home, Community centers, Medical specialty clinics, Nursing Homes, Parks, Primary care |
| Delivering Entity | The organization or person delivering the intervention |  |
| Intervention Type | The intervention being studied | Qi Gong, Tai Chi, Yoga |
| Strength of Intervention Evidence | The strength of the evidence for an intervention in a clinical population as referenced or justified by the authors | Guideline recommending intervention, guideline recommending component of intervention (e.g., exercise, mindfulness), other evidence |
| Intervention Mode | The delivery of the intervention | In-person groups, Virtual groups, In-person individual, Virtual individual |
| Intervention Duration | The duration of the intervention |  |
| Intervention Frequency | The frequency of the intervention |  |
| Clinical Population/Health Condition | The population this study would be applied/generalized to |  |
| Other Participant Characteristics | Other defining characteristics of the sample |  |
| Sample Size | Number of people in the sample |  |
| Mean Age | Average age of the sample |  |
| % Female | Percentage of female participants in the sample |  |
| % Non-white | Percentage of non-white participants in the sample |  |
| Implementation Framework | Implementation framework used in the study | Yes or no |
| Implementation Outcome(s) | Identify the implementation outcome of interest if they used one | Acceptability, Adoption, Appropriateness, Cost, Feasibility, Fidelity, Penetration, Sustainability |
| CFIR Domain – Innovation | Extract text/content from the results section of article related to one or more of the constructs from the innovation domain | Code each distinct statement with CFIR construct and whether it was a barrier, facilitator, both, or neither |
| CFIR Domain – Outer Setting | Extract text/content from the results section of article related to one or more of the constructs from the outer setting domain | Separate distinct statements and code with individual CFIR constructs. Indicate whether each construct represents a barrier, facilitator, both, or neither |
| CFIR Domain – Inner Setting | Extract text/content from the results section of article related to one or more of the constructs from the inner setting domain | Separate distinct statements and code with individual CFIR constructs. Indicate whether each construct represents a barrier, facilitator, both, or neither |
| CFIR Domain – Individuals | Extract text/content from the results section of article related to one or more of the constructs from the individuals domain | Separate distinct statements and code with individual CFIR constructs. Indicate whether each construct represents a barrier, facilitator, both, or neither |
| CFIR Domain – Process | Extract text/content from the results section of article related to one or more of the constructs from the process domain | Separate distinct statements and code with individual CFIR constructs. Indicate whether each construct represents a barrier, facilitator, both, or neither |
| <sup>a</sup>Extraction of initial articles will inform refinement of the extraction guide and further development of study characteristics, their definitions, and categories for each characteristic. This will be an iterative process, and the extraction form will be updated as necessary, through discussion at weekly team meetings. A final extraction form will be published with our main findings. |  |  |

